# Interim Analysis of a Phase I Randomized Clinical Trial on the Safety and Immunogenicity of the mRNA-1283 SARS-CoV-2 Vaccine in Adults

**DOI:** 10.1101/2022.10.18.22281050

**Authors:** Patrick Yassini, Mark Hutchens, Yamuna D. Paila, Lorraine Schoch, Anne Aunins, Uma Siangphoe, Robert Paris

**Author notes:** **Corresponding Author:** Robert Paris, MD, Head, Translational Medicine, Infectious Disease Development, Moderna, Inc., 200 Technology Square, Cambridge, MA 02139. **Alternative Corresponding Author:** Anne Aunins, Program Leader, COVID-19 Vaccines, Moderna, Inc., 200 Technology Square, Cambridge, MA 02139.

## Abstract

**Background:** This interim analysis of an ongoing phase I randomized clinical trial evaluated the safety, reactogenicity, and immunogenicity of mRNA-1283, a next-generation SARS-CoV-2 messenger RNA (mRNA)-based vaccine encoding 2 segments of the spike protein (ie, receptor binding and N-terminal domains).

**Methods:** Healthy aged adults 18-55 years (n = 104) were randomized (1:1:1:1:1) to receive 2 doses of mRNA-1283 (10, 30, or 100 μg) or mRNA-1273 (100 μg) administered 28 days apart, or a single dose of mRNA-1283 (100 μg). Safety was assessed and immunogenicity was measured by serum neutralizing antibody (nAb) or binding antibody (bAb) responses.

**Results:** At the interim analysis, no safety concerns were identified and no serious adverse events, adverse events of special interest, or deaths were reported. Solicited systemic adverse reactions were more frequent with higher dose levels of mRNA-1283 than with mRNA-1273. At day 57, all dose levels of the 2-dose mRNA-1283 regimen (including the lowest dose level [10 μg]) induced robust nAb and bAb responses that were comparable to those of mRNA-1273 (100 μg).

**Conclusions:** mRNA-1283 was generally safe in adults, with all dose levels of the 2-dose regimen (10, 30, and 100 μg) eliciting similar immunogenicity as the 2-dose mRNA-1273 regimen (100 μg).

**Clinical Trials Registration:** Clinicaltrials.gov, NCT04813796

## INTRODUCTION

An unprecedented global response to the coronavirus disease 2019 (COVID-19) pandemic led to the rapid development of vaccines against the causative pathogen, severe acute respiratory syndrome coronavirus-2 (SARS-CoV-2). The messenger RNA (mRNA)-based vaccine mRNA-1273 (SPIKEVAX; Moderna, Inc., Cambridge, MA, USA [1]) is authorized and approved for use in many populations worldwide, available for individuals as young as ≥6 months [2]. In the phase III clinical trial in adults, mRNA-1273 had an acceptable safety profile and was efficacious against symptomatic infection and severe disease [3]. Subsequent post-authorization real-world studies provided evidence of mRNA-1273 effectiveness against symptomatic infection and hospitalization [4-6] but demonstrated reduced effectiveness against infection caused by several SARS-CoV-2 variants of concern, including B.1.351 (beta), AY lineages (delta), and BA lineages (omicron) [7-9]. A booster dose is recommended in individuals aged ≥12 years to maintain effectiveness [10].

The response to the COVID-19 pandemic demonstrated that the mRNA-lipid nanoparticle (LNP) platform affords several advantages, including expeditious development of safe and efficacious vaccines [11]; however, the refrigerated shelf life of some of these vaccines can limit vaccine distribution in certain markets [12-15]. Next-generation mRNA-based SARS-CoV-2 vaccines that offer longer refrigerated shelf life and/or are immunogenic at a lower dose than mRNA-1273 may enable increased mRNA-based vaccine coverage worldwide, potentially further preventing COVID-19–associated morbidity and mortality.

mRNA-1283 is one such investigational vaccine candidate, aiming to elicit neutralizing antibodies by encoding 2 segments of the SARS-CoV-2 S protein, including the receptor binding domain (RBD) and the N-terminal domain (NTD) regions, which contain key SARS-CoV-2 neutralization epitopes [16, 17]. Comparatively, mRNA-1273 encodes the full-length SARS-CoV-2 S protein. A preclinical evaluation of mRNA-1283 in mice demonstrated comparable protection and immunogenicity characteristics as mRNA-1273 with potential dose-sparing properties, while indicating that the shorter mRNA length within mRNA-1283 may facilitate longer storage under refrigeration than mRNA-1273 [18].

Here, we report preliminary safety, reactogenicity, and immunogenicity results for mRNA-1283 from a phase I, randomized, observer-blind, dose-ranging study in healthy adults aged 18-55 years.

## METHODS

### Trial Design and Participants

This is an ongoing phase I, randomized, observer-blind study being conducted at 5 sites in the United States to evaluate the safety, reactogenicity, and immunogenicity of mRNA-1283 (NCT04813796). Eligible participants were healthy adults (aged 18-55 years) with negative SARS-CoV-2 serology, no known history of SARS-CoV-2 infection or exposure to SARS-CoV-2 or COVID-19 in the previous 30 days, and were not previously administered any coronavirus vaccine. A full list of the study inclusion/exclusion criteria is in the **Supplement**.

Three dose levels of mRNA-1283 (10 μg, 30 μg, and 100 μg) and 1 dose level of mRNA-1273 (100 μg) were administered on a 2-dose schedule with 28 days between doses; the mRNA-1283 100-μg dose level was also assessed as a single-dose regimen. Participants were randomly assigned in parallel (1:1:1:1:1) using interactive response technology (**Figure S1**; see the **Supplement** for details). Participants were followed for safety, reactogenicity, and immunogenicity, with a planned interim analysis to evaluate data on ≥80 participants through day 57. After the interim analysis, participants who received the single mRNA-1283 100-μg dose were offered an additional injection of mRNA-1273 (100 μg).

The protocol was approved by the central institutional review board (Advarra, Inc; Columbia, MD) and the study was conducted in accordance with the protocol, applicable laws and regulatory requirements, the International Council for Harmonization Good Clinical Practice guidelines, and the ethical principles derived from the Declaration of Helsinki and Council for International Organizations of Medical Sciences International Ethical Guidelines. All participants provided written informed consent before any study procedures were conducted.

### Vaccines

mRNA-1283 is an mRNA-LNP vaccine encoding for a chimeric protein (NTD-RBD-HATM) comprising the SARS-CoV-2 S protein NTD and RBD, linked together by a flexible peptide linker and anchored to a 23-amino acid transmembrane domain from influenza hemagglutinin (HATM). mRNA-1273 is an mRNA-LNP vaccine encoding for the full-length, 2-proline prefusion stabilized SARS-CoV-2 S protein (S2P). Placebo was a 0.9% sodium chloride (normal saline) injection.

Vaccines were provided as a sterile liquid for injection at 0.2 mg/mL (mRNA-1273) or 0.5 mg/mL (mRNA-1283; diluted with 0.9% saline to the appropriate dose) concentrations; all vaccines were administered at 0.5-mL dose volume into the deltoid muscle. For the 2-dose regimen, each dose level of mRNA-1283 or mRNA-1273 was administered 28 days apart (day 1 and day 29). The 1-dose regimen of mRNA-1283 had placebo administered on day 1 and mRNA-1283 (100 μg) administered on day 29.

### Objectives and Endpoints

The primary objective was to evaluate the safety and reactogenicity of mRNA-1283 (10 μg, 30 μg, and 100 μg) and mRNA-1273 (100 μg), each administered on a 2-dose schedule (28 days apart), and mRNA-1283 (100 μg) administered as a single dose. Safety endpoints included solicited local (ie, pain, erythema, or hardness at the injection site; or axillary swelling or tenderness) and systemic (ie, headache, fatigue, myalgia, arthralgia, nausea, chills, and fever) adverse reactions (ARs) ≤7 days of each dose, as recorded by participants using an electronic diary; safety laboratory abnormalities ≤7 days of each dose (see the **Supplement**); unsolicited adverse events (AEs) ≤28 days of each dose; and any serious AEs (SAEs), medically attended AEs (MAAEs), AEs leading to discontinuation, and AEs of special interest (AESIs) from day 1 to end of study. Safety data were monitored by an unblinded, independent data and safety monitoring board, a blinded internal safety team, and a blinded safety oversight team. This report summarizes interim safety assessments after a median of 127 days (>4 months) after dose 2.

Secondary objectives were to evaluate the immunogenicity of mRNA-1283 administered as a 2-dose schedule (10 μg, 30 μg, and 100 μg) or a single dose schedule (100 μg) and mRNA-1273 administered as a 2-dose schedule (100 μg) as measured by nAb titer or level of binding antibody (bAb). Serum nAb titers were measured by a validated lentivirus assay [19, 20] (PsVN assay; see the **Supplement**) against Wuhan-Hu-1 (with D614G mutation) SARS-CoV-2 or the B.1.351 (beta) variant with a 50% inhibitory dilution; titers against Wuhan-Hu-1 (without D614G mutation) were also measured by a validated live virus 50% microneutralization assay [21] (see the **Supplement**). Levels of serum bAbs against Wuhan-Hu-1 SARS-CoV-2 were measured by a SARS-CoV-2 Meso Scale Discovery (MSD) 4-PLEX assay (modified 3-PLEX assay [22]; see the **Supplement**). For this interim analysis, only assessments on samples collected on day 1 (baseline) and days 29, 36, and 57 were summarized.

### Statistical Analyses

Sample size determination is described in the **Supplement**. Safety was assessed in the safety and solicited safety populations (defined in the **Supplement**). All safety analyses were summarized according to the actual vaccine received. Safety was descriptively analyzed as counts and percentages.

Immunogenicity was assessed in the per-protocol population (defined in the **Supplement**). For each vaccine group, the geometric mean titer (GMT) or levels of SARS-CoV-2 nAb and bAb were computed at each sampling time point, as well as the geometric mean fold rise (GMFR) of titers or levels from baseline to each post-baseline time point; values were summarized alongside corresponding 95% confidence intervals (CIs) calculated based on the *t* distribution of the log-transformed values, then back-transformed to the original scale for presentation. The geometric mean ratio (GMR) calculated as the mean of the difference in nAb titers and bAb levels between mRNA-1283 and mRNA-1273 and the corresponding 2-sided 95% CI were provided. Percentages of participants with vaccine seroresponse were also calculated by time point and vaccine group alongside 2-sided 95% CIs using the Clopper-Pearson method. Seroresponse was defined as an increase in titer or level from less than the lower limit of quantification [LLOQ] at baseline to ≥LLOQ, or a ≥4-fold increase over baseline in participants with pre-existing baseline titers or levels that were ≥LLOQ. All analyses were performed using SAS software, version 9.4 (SAS Institute).

## RESULTS

### Participants

Between March 11, 2021, and July 28, 2021, a total of 104 participants were randomly assigned to receive 2 doses of mRNA-1283 (10 μg, n=21; 30 μg, n=22; 100 μg, n=21), 1 dose of mRNA-1283 (100 μg; n=18), or 2 doses of mRNA-1273 100 μg (n=22). All randomly assigned participants received dose 1 and 75 (91.5%) and 20 participants (90.9%) received dose 2 in the mRNA-1283 and mRNA-1273 groups, respectively (**Figure 1**). Overall, 68 participants (82.9%) in the mRNA-1283 groups and 18 participants (81.8%) in the mRNA-1273 group were included in the per-protocol immunogenicity population for this interim analysis. Participant baseline demographics are summarized in **Table 1**.

**Table 1.**
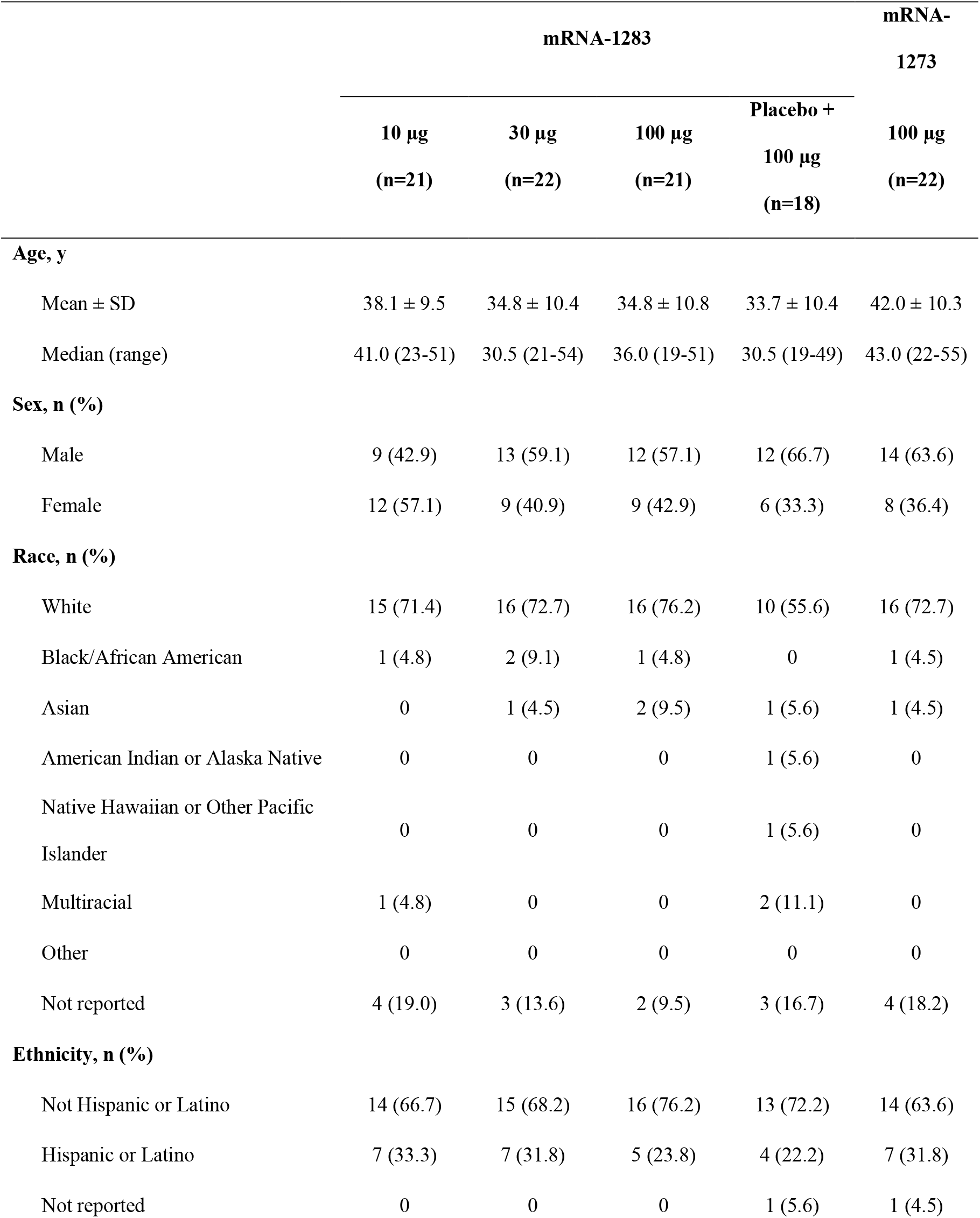
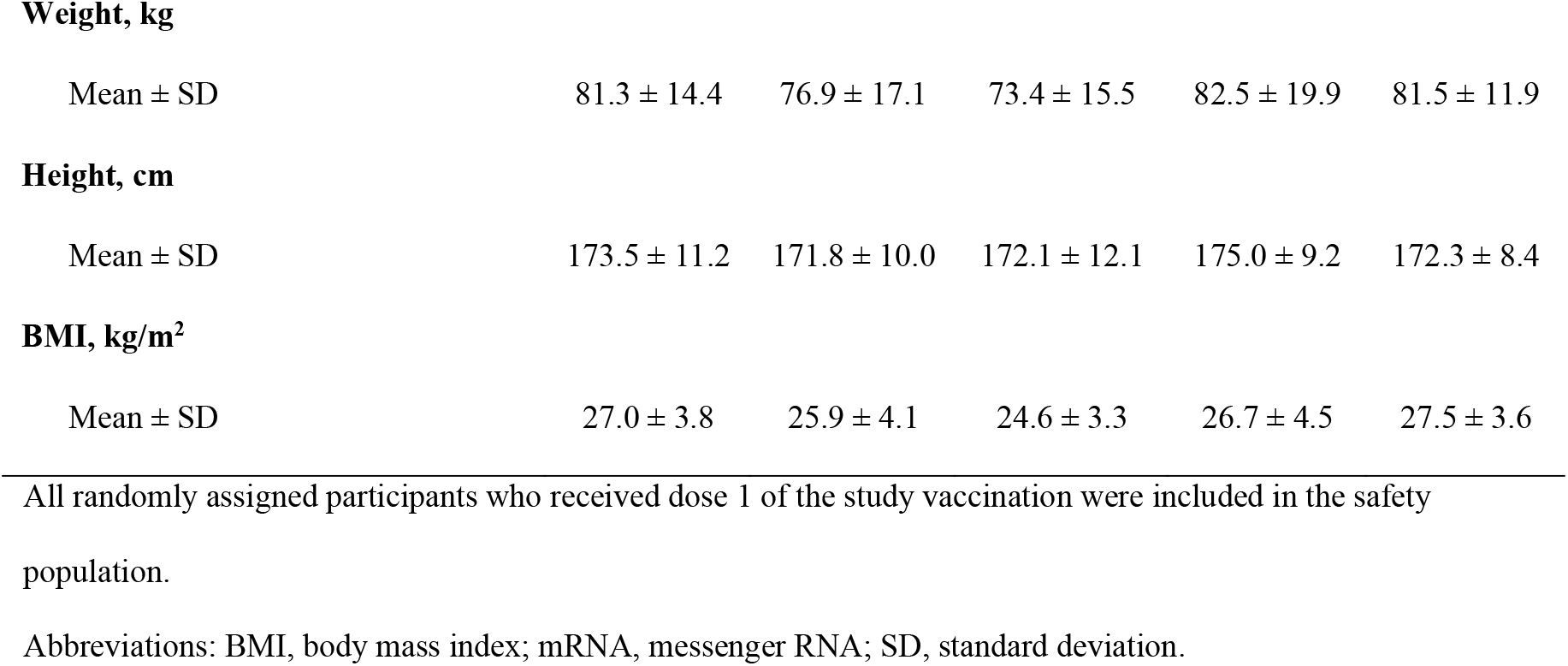
Participant baseline demographics (safety population)

|  | mRNA-1283 |  |  |  | mRNA-1273 |
| --- | --- | --- | --- | --- | --- |
|  | 10 µg<br>(n=21) | 30 µg<br>(n=22) | 100 µg<br>(n=21) | Placebo +<br>100 µg<br>(n=18) | 100 µg<br>(n=22) |
| **Age, y** |  |  |  |  |  |
| Mean ± SD | 38.1 ± 9.5 | 34.8 ± 10.4 | 34.8 ± 10.8 | 33.7 ± 10.4 | 42.0 ± 10.3 |
| Median (range) | 41.0 (23-51) | 30.5 (21-54) | 36.0 (19-51) | 30.5 (19-49) | 43.0 (22-55) |
| **Sex, n (%)** |  |  |  |  |  |
| Male | 9 (42.9) | 13 (59.1) | 12 (57.1) | 12 (66.7) | 14 (63.6) |
| Female | 12 (57.1) | 9 (40.9) | 9 (42.9) | 6 (33.3) | 8 (36.4) |
| **Race, n (%)** |  |  |  |  |  |
| White | 15 (71.4) | 16 (72.7) | 16 (76.2) | 10 (55.6) | 16 (72.7) |
| Black/African American | 1 (4.8) | 2 (9.1) | 1 (4.8) | 0 | 1 (4.5) |
| Asian | 0 | 1 (4.5) | 2 (9.5) | 1 (5.6) | 1 (4.5) |
| American Indian or Alaska Native | 0 | 0 | 0 | 1 (5.6) | 0 |
| Native Hawaiian or Other Pacific Islander | 0 | 0 | 0 | 1 (5.6) | 0 |
| Multiracial | 1 (4.8) | 0 | 0 | 2 (11.1) | 0 |
| Other | 0 | 0 | 0 | 0 | 0 |
| Not reported | 4 (19.0) | 3 (13.6) | 2 (9.5) | 3 (16.7) | 4 (18.2) |
| **Ethnicity, n (%)** |  |  |  |  |  |
| Not Hispanic or Latino | 14 (66.7) | 15 (68.2) | 16 (76.2) | 13 (72.2) | 14 (63.6) |
| Hispanic or Latino | 7 (33.3) | 7 (31.8) | 5 (23.8) | 4 (22.2) | 7 (31.8) |
| Not reported | 0 | 0 | 0 | 1 (5.6) | 1 (4.5) |
| **Weight, kg** |  |  |  |  |  |
| Mean ± SD | 81.3 ± 14.4 | 76.9 ± 17.1 | 73.4 ± 15.5 | 82.5 ± 19.9 | 81.5 ± 11.9 |
| **Height, cm** |  |  |  |  |  |
| Mean ± SD | 173.5 ± 11.2 | 171.8 ± 10.0 | 172.1 ± 12.1 | 175.0 ± 9.2 | 172.3 ± 8.4 |

**BMI, kg/m<sup>2</sup>**

|  |  |  |  |  |  |
| --- | --- | --- | --- | --- | --- |
| Mean ± SD | 27.0 ± 3.8 | 25.9 ± 4.1 | 24.6 ± 3.3 | 26.7 ± 4.5 | 27.5 ± 3.6 |

Abbreviations: BMI, body mass index; mRNA, messenger RNA; SD, standard deviation.

**Figure 1.**
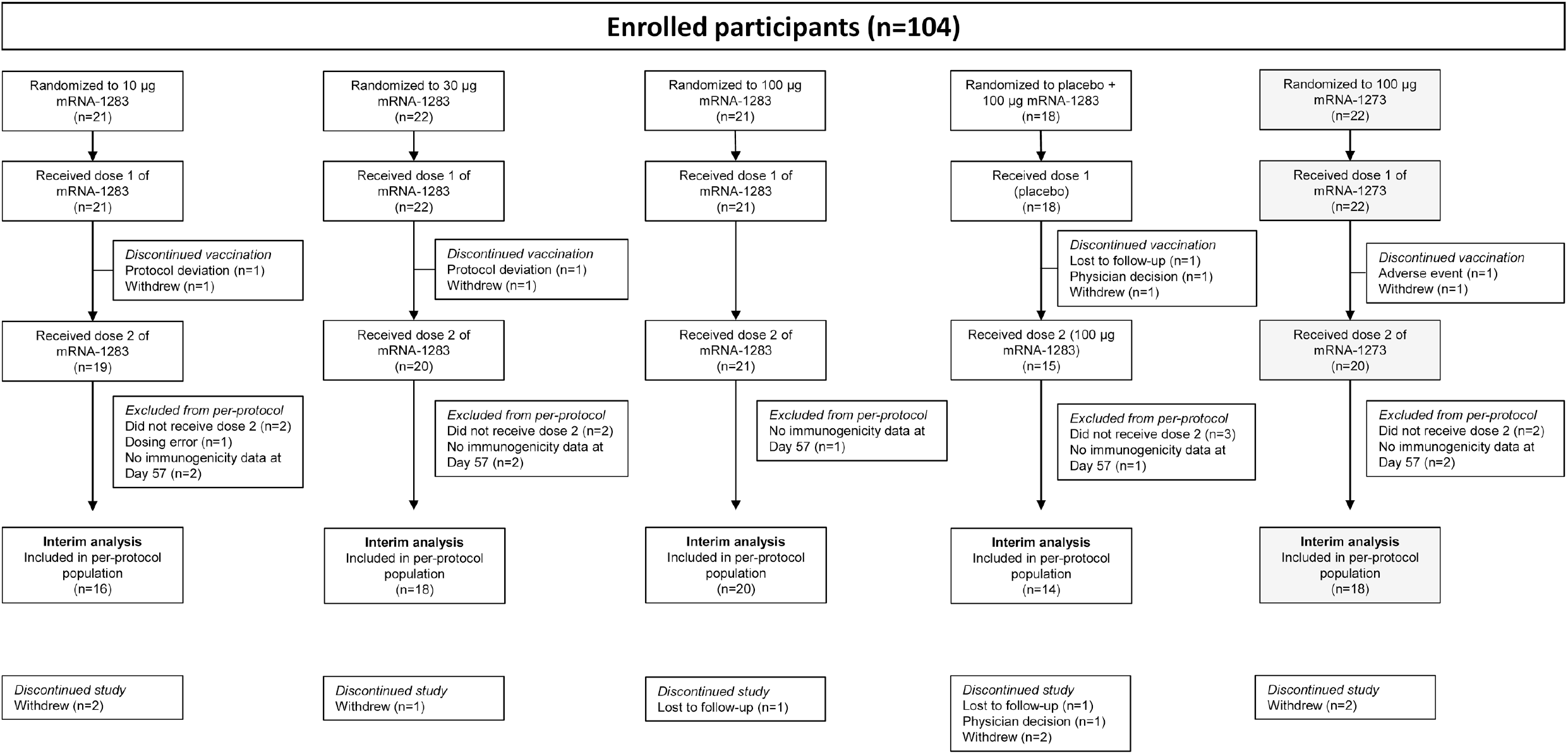
Participant disposition. One participant in the mRNA-1273 100-μg group discontinued vaccination because of an adverse event, which was considered related to study vaccination. All randomly assigned participants who received dose 1 of the study vaccination were included in the safety population. Reasons for exclusion from the per-protocol immunogenicity population are described in the **Supplementary Materials**. Abbreviation: mRNA, messenger RNA.

### Safety

#### Solicited Local and Systemic Adverse Reactions

Across mRNA-1283 groups, solicited local ARs ≤7 days of vaccination were reported by 6-18 participants (33.3-85.7%) after dose 1 and 12-19 participants (73.7-90.5%) after dose 2 (**Figure 2A**); 21 (95.5%) and 19 mRNA-1273 recipients (95.0%) reported solicited local ARs after dose 1 and dose 2, respectively. The majority of solicited local ARs were mild to moderate (grades 1 and 2), with no grade 4 events reported. Pain at the injection site was most common, reported by >70% of participants across vaccine groups following dose 2. The median duration of solicited local ARs was 2-3 days after mRNA-1283 and 3 days after mRNA-1273.

**Figure 2.**
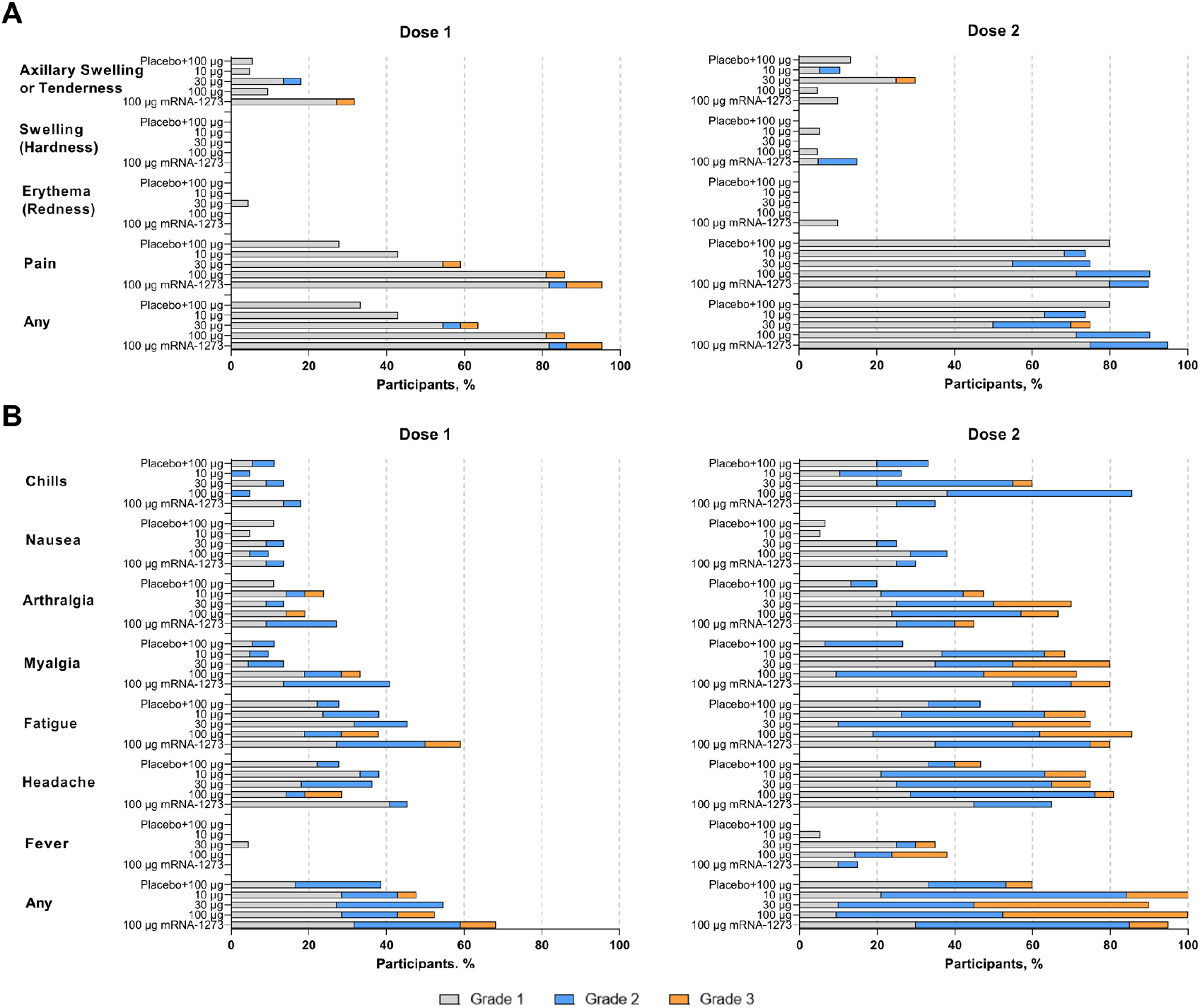
Solicited (A) local and (B) systemic adverse reactions within 7 days of each dose. Percentages are based on participants in the solicited safety population, consisting of all randomly assigned participants receiving ≥1 dose and contributing any solicited adverse reaction data. Number of participants in the dose 1 solicited safety population: mRNA-1283 10 μg, n=21; mRNA-1283 30 μg, n=22; mRNA-1283 100 μg, n=21; placebo + mRNA-1283 100 μg, n=18; and mRNA-1273 100 μg, n=22. Number of participants in the dose 2 solicited safety population: mRNA-1283 10 μg, n=19; mRNA-1283 30 μg, n=20; mRNA-1283 100 μg, n=21; placebo + mRNA-1283 100 μg, n=15; and mRNA-1273 100 μg, n=20. Abbreviation: mRNA, messenger RNA.

After dose 1, solicited systemic ARs ≤7 days of vaccination were reported by 7-12 participants (38.9-54.5%) across mRNA-1283 groups and 15 participants (68.2%) in the mRNA-1273 group (**Figure 2B**). Systemic ARs were more frequent after dose 2, reported by 9-21 participants (60-100%) across mRNA-1283 groups and 19 participants (95.0%) in the mRNA-1273 group. Headache, fatigue, and myalgia were most frequent. Solicited systemic ARs were predominantly mild to moderate after dose 1 of mRNA-1283, but increased in severity after dose 2, with 3 to 12 (20.0-63.2%) and 1 to 10 participants (6.7-47.6%) reporting grade 2 and 3 events, respectively; no grade 4 events were reported. Increased severity was predominantly reported at higher mRNA-1283 dose levels. Fever and chills were largely reported after dose 2 and for participants who received mRNA-1283 at higher dose levels. The median duration of solicited systemic ARs was 2-3 days after mRNA-1283 and 2 days after mRNA-1273.

#### Unsolicited Adverse Events

No prespecified study pause rules were met, and no SAEs, AESIs, severe AEs, or fatal AEs were reported ≤28 days after any dose of mRNA-1283 or mRNA-1273. Unsolicited treatment-emergent AEs (TEAEs) were reported by 4-9 participants (22.2-42.9%) across mRNA-1283 groups and 7 participants (31.8%) in the mRNA-1273 group (**Table 2**). Overall, 2-4 (9.5-19.0%) and 4 participants (18.2%) reported unsolicited TEAEs considered related to either mRNA-1283 or mRNA-1273, respectively. Across mRNA-1283 groups, a total of 2-3 participants (9.1-14.3%) reported MAAEs; 2 participants reported vaccine-related MAAES (n=1, allergic dermatitis [mRNA-1283 30 μg]; n=1, headache, fatigue, arthralgia, and myalgia [mRNA-1283 100 μg]). Three mRNA-1273 recipients (13.6%) reported MAAEs, and 1 MAAE (vomiting, 4.5%) was considered related to vaccination. One participant in the mRNA-1273 group discontinued vaccination because of transient elevation in liver transaminase levels 7 days after dose 1 that subsequently returned to normal, which was considered vaccine-related. Clinical laboratory findings are presented in the **Supplement**.

**Table 2.** Summary of unsolicited TEAEs through 28 days following any vaccination (safety population)

|  | mRNA-1283 |  |  |  | mRNA-1273 |
| --- | --- | --- | --- | --- | --- |
|  | 10 µg<br>(n=21) | 30 µg<br>(n=22) | 100 µg<br>(n=21) | Placebo +<br>100 µg<br>(n=18) | 100 µg<br>(n=22) |
| n (%) |  |  |  |  |  |
| **All unsolicited TEAEs** |  |  |  |  |  |
| All AEs | 8 (38.1) | 9 (40.9) | 9 (42.9) | 4 (22.2) | 7 (31.8) |
| SAEs | 0 | 0 | 0 | 0 | 0 |
| Fatal AEs | 0 | 0 | 0 | 0 | 0 |
| MAAEs | 3 (14.3) | 2 (9.1) | 2 (9.5) | 2 (11.1) | 3 (13.6) |
| AEs leading to vaccination<br>discontinuation | 0 | 0 | 0 | 0 | 1 (4.5) |
| AEs leading to study<br>discontinuation | 0 | 0 | 0 | 0 | 0 |
| Severe | 0 | 0 | 0 | 0 | 0 |
| AESIs | 0 | 0 | 0 | 0 | 0 |
| **Related unsolicited TEAEs** |  |  |  |  |  |
| All AEs | 2 (9.5) | 4 (18.2) | 4 (19.0) | 2 (11.1) | 4 (18.2) |
| SAEs | 0 | 0 | 0 | 0 | 0 |
| Fatal AEs | 0 | 0 | 0 | 0 | 0 |
| MAAEs | 0 | 1 (4.5) | 1 (4.8) | 0 | 1 (4.5) |
| AEs leading to vaccination<br>discontinuation | 0 | 0 | 0 | 0 | 1 (4.5) |
| AEs leading to study<br>discontinuation | 0 | 0 | 0 | 0 | 0 |
| Severe | 0 | 0 | 0 | 0 | 0 |
| AESIs | 0 | 0 | 0 | 0 | 0 |

### Immunogenicity

#### Serum Neutralizing Antibodies Against SARS-CoV-2

For the 2-dose regimen, pseudovirus nAb GMTs to D614G SARS-CoV-2 increased from baseline after dose 1 (day 29) of mRNA-1273 (100 μg) and all dose levels of mRNA-1283 (10 μg, 30 μg, and 100 μg; **Figure 3A**). At 28 days following dose 2 (day 57), pseudovirus nAb GMTs (95% CI) for all dose levels of mRNA-1283 administered were comparable to those of mRNA-1273 (mRNA-1283, 10 μg: 1370 [888-2112], 30 μg: 1091 [645-1846], and 100 μg: 1679 [1133-2487]; mRNA-1273: 1190 [826-1713]; **Figure 3A**; **Table S1**); generally similar finding were observed against Wuhan-Hu-1 (without D614G mutation) with the live-virus microneutralization assay (**Figure S2**; **Table S1**). As measured by pseudovirus nAb titers against D614G, seroresponse at day 57 occurred in 100% of participants in all groups who received 2 doses of mRNA vaccines. Geometric mean ratios (95% CI; **Table S1**) of GMTs for 2-dose regimens of mRNA-1283 versus mRNA-1273 at day 57 were 1.15 (0.67-1.98), 0.92 (0.50-1.70), and 1.41 (0.84-2.38) for the mRNA-1283 10-μg, 30-μg, and 100-μg dose levels, respectively. For the 1-dose mRNA-1283 regimen (placebo + mRNA-1283 100 μg), a slight increase from baseline was observed 28 days after mRNA-1283 administration (day 57) but remained lower than 2-dose regimens; the GMR (95% CI) at day 57 relative to 2 doses of mRNA-1273 was 0.05 (0.01-0.18) and seroresponse occurred in 28.6% of participants.

**Figure 3.**
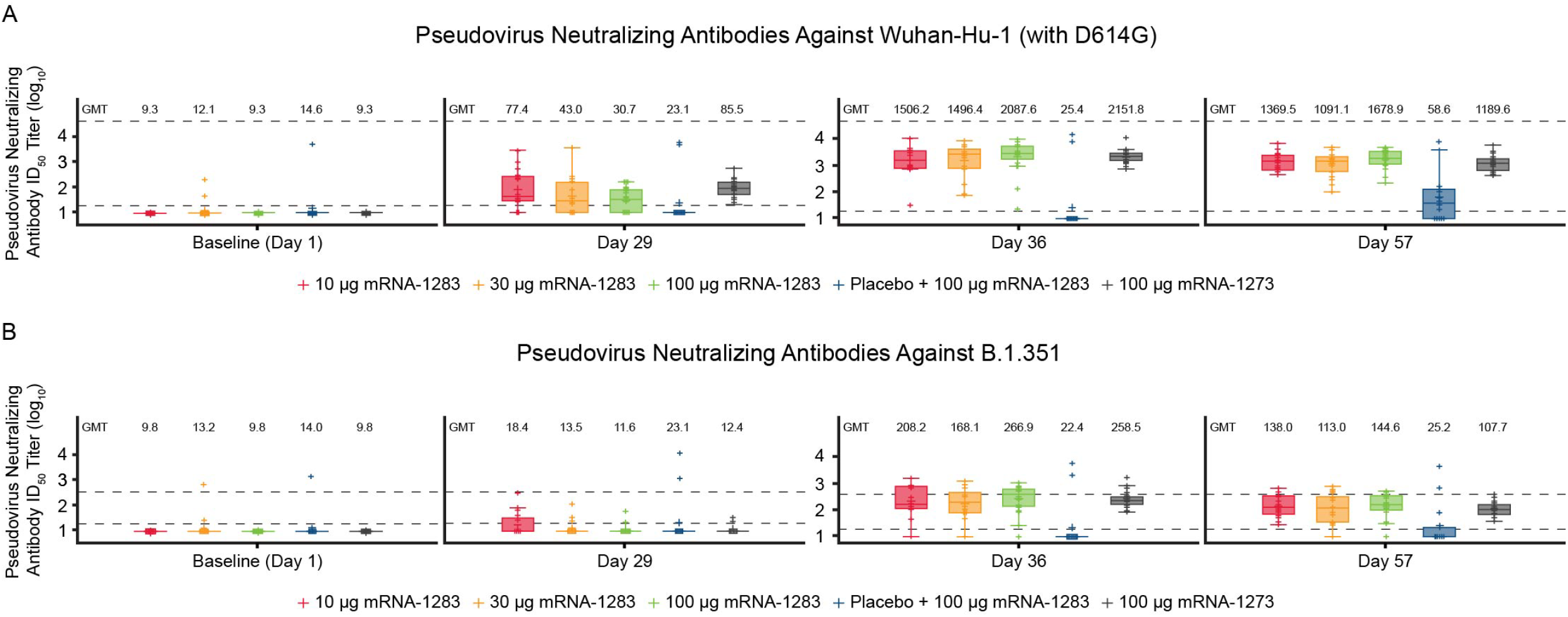
Pseudovirus neutralizing antibody ID_50_ titers against (A) Wuhan-Hu-1 (with D614G mutation) SARS-CoV-2 or (B) B.1.351 variant through day 57. Neutralizing antibody titers on the pseudovirus neutralization assay at ID_50_ (log scale) are shown for those participants in the per-protocol immunogenicity population. In both panels, boxplots are based on log-transformed values; boxes and horizontal bars denote IQR range and ID_50_, respectively, with whisker end points equal to the values below or above the median at 1.5 times the IQR. Dots represent individual results and the dotted lines represent either the LLOQ or ULOQ. Antibody values reported as below the LLOQ were replaced by 0.5 x LLOQ, whereas values greater than the ULOQ were converted to the ULOQ if the actual values were unavailable. In panel A, the LLOQ was 18.5 (log_10_[LLOQ] = 1.267) and the ULOQ was 45118 (log_10_[ULOQ] = 4.654). In panel B, the LLOQ was 19.5 (log_10_[LLOQ] = 1.290) and the ULOQ was 385.7 (log_10_[ULOQ] = 2.586). Abbreviations: ID_50_, 50% inhibitory dose; IQR, interquartile range; LLOQ, lower limit of quantification; mRNA, messenger RNA; SARS-CoV-2, severe acute respiratory syndrome coronavirus-2; ULOQ, upper limit of quantification.

After dose 2, all 2-dose mRNA-1283 groups (10 μg, 30 μg, and 100 μg) had comparable pseudovirus nAb GMTs to B.1.351 compared with mRNA-1273 (**Figure 3B; Table S1**). The 1-dose mRNA-1283 100 μg regimen did not induce a substantial nAb response to B.1.351 at day 57 (**Figure 3B**). At day 57, seroresponse to B.1.351 occurred in 61.1%-85.0% and 66.7% of participants in the 2-dose mRNA-1283 groups and mRNA-1273 group, respectively (**Table S1**); 14.3% of participants in the 1-dose mRNA-1283 regimen had a seroresponse to B.1.351.

#### Serum Binding Antibodies Against SARS-CoV-2

For the 2-dose regimens, bAb immunoglobulin G (IgG) responses to Wuhan-Hu-1 full-length S protein were low or undetectable at baseline for all vaccine groups and increased after dose 1 of mRNA-1283 (10 μg, 30 μg, and 100 μg) and mRNA-1273 (100 μg; **Figure 4A**). At 28 days after dose 2 (day 57), bAb responses were generally comparable between mRNA-1283 and mRNA-1273 regardless of dose level (**Figure 4A**; **Table S2**), with GM levels (95% CI) reported as 322,671 (232,477-447,858), 280,454 (196,077-401,139), 416,620 (312,082-556,175), and 303,004 (242,750-378,212) for the mRNA-1283 10-μg, 30-μg, and 100-μg and mRNA-1273 100-μg dose levels, respectively. Seroresponse at day 57 occurred in 100% of mRNA-1283 (all dose levels) or mRNA-1273 100 μg recipients. Similar trends were observed for bAb IgG responses to Wuhan-Hu-1 S protein RBD and NTD (**Figure 4B-C**; **Table S2**), with all dose levels of mRNA-1283 eliciting similar responses to either domain at day 57 as compared with mRNA-1273; seroresponses were observed in 100% of 2-dose vaccine recipients at day 57.

**Figure 4.**
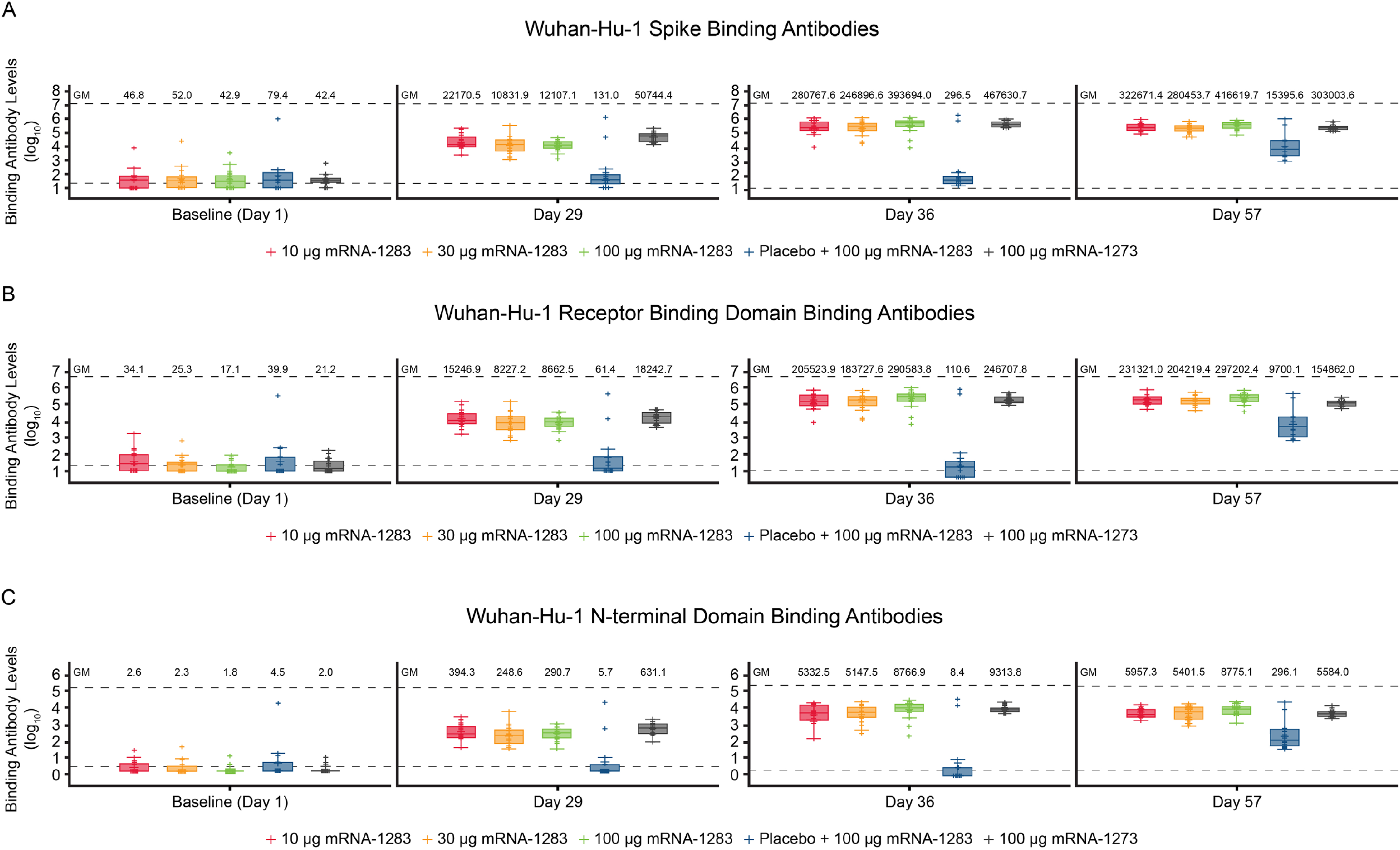
Binding antibody levels through day 57 specific to Wuhan-Hu-1 SARS-CoV-2 (A) S protein or (B) RBD. Binding IgG antibody levels specific to Wuhan-Hu-1 SARS-CoV-2 S protein (panel A), RBD (panel B), or NTD (panel C) by MSD 4-PLEX assay are shown for those participants in the per-protocol immunogenicity population. In both panels, boxplots are based on log-transformed values; boxes and horizontal bars denote IQR range and GM, respectively, with whisker end points equal to the values below or above the median at 1.5 times the IQR. Individual results are represented as dots and the dotted lines represent either the LLOQ or ULOQ. Antibody values reported as below the LLOQ were replaced by 0.5 x LLOQ, whereas values greater than the ULOQ were converted to the ULOQ if actual values were unavailable. In panel A, the LLOQ was 23 (log_10_[LLOQ] = 1.362) and the ULOQ was 14000000 (log_10_[ULOQ] = 7.146). In panel B, the LLOQ was 19 (log_10_[LLOQ] = 1.279) and the ULOQ was 6000000 (log_10_[ULOQ] = 6.778). Abbreviations: ID_50_, 50% inhibitory dose; IgG, immunoglobulin G; IQR, interquartile range; LLOQ, lower limit of quantification; mRNA, messenger RNA; MSD, Meso Scale Discovery; NTD, N-terminal domain; RBD, receptor binding domain; SARS-CoV-2, severe acute respiratory syndrome coronavirus-2; ULOQ, upper limit of quantification.

The 1-dose mRNA-1283 regimen (placebo + mRNA-1283 100 μg) showed an increase from baseline in bAb responses to Wuhan-Hu-1 S protein, RBD, and NTD at 28 days after mRNA-1283 was administered (day 57). GM levels (95% CI) were 15,396 (4855-48,820) for S protein, 9700 (3110-30,252) for RBD, and 296 (95-919) for NTD; seroresponse against S protein, RBD, and NTD was 92.9%.

## DISCUSSION

Here, we presented interim safety and immunogenicity findings of a phase I clinical study in healthy adults of a next-generation SARS-CoV-2 vaccine candidate (mRNA-1283), which encodes the SARS-CoV-2 S protein RBD and NTD segments versus full-length S protein encoded by mRNA-1273. Preliminary findings indicate that all mRNA-1283 dose levels (10 μg, 30 μg, and 100 μg) administered on a 2-dose schedule (28 days apart) were generally safe and elicited robust immune responses that were comparable to mRNA-1273 (100 μg). The lowest investigated mRNA-1283 dose level (10 μg) had the most favorable tolerability profile and induced robust nAb and bAb responses to SARS-CoV-2 after dose 2 that were similar to those of mRNA-1273. Overall, these findings suggest that a 2-dose regimen of mRNA-1283 at a lower dose level may achieve similar vaccine efficacy to mRNA-1273.

No safety concerns were identified and no deaths, SAEs, or AESIs were reported. Both local and systemic ARs after dose 1 of mRNA-1283 were generally mild to moderate, characterized by pain at the injection site, headache, fatigue, and myalgia. After dose 2, solicited systemic ARs generally increased in frequency and severity. Overall, compared with mRNA-1273, solicited systemic ARs occurred more frequently with higher dose levels of mRNA-1283.

Following a 2-dose schedule, all mRNA-1283 dose levels (including the lowest dose [10 μg]) induced nAb responses after dose 2 to prototype SARS-CoV-2 that were comparable to mRNA-1273 (100 μg). Notably, the pseudovirus nAb responses after dose 2 of mRNA-1273 in this study align with those among adults in the pivotal phase III trial [23], wherein 2 doses of mRNA-1273 (100 μg) administered 28 days apart were efficacious against symptomatic COVID-19 infection and severe COVID-19 [3, 24]. Additionally, levels of bAb responses to full-length S protein, NTD, and RBD were similar between mRNA-1273 and all mRNA-1283 dose levels at 28 days following dose 2. Our interim phase I findings raise the possibility that a lower mRNA-1283 dose administered via a 2-dose regimen might achieve similar efficacy and effectiveness as mRNA-1273 among adults. Notably, although the 1-dose regimen of mRNA-1283 (100 μg) induced bAb responses at 28 days, levels remained lower than those for 2-dose mRNA-1283 and mRNA-1273 regimens; further, neutralizing responses to D614G SARS-CoV-2 remained low after 1 dose, together supporting a 2-dose schedule of mRNA-1283. These data suggest that higher mRNA-1283 dose levels do not result in higher nAb responses; however, whether this finding extends to T-cell responses is currently being assessed. Because the severity of solicited systemic ARs was increased at the mRNA-1283 100-μg dose level, it remains possible that increased systemic inflammation at higher dose levels, potentially through complex interactions of type 1 interferons or other innate immune effector molecules, may suppress B- and/or T-cell responses and warrants further study [25-27]. The 2-dose schedule of mRNA-1283 (all dose levels) and mRNA-1273 also elicited comparable nAb responses to the SARS-CoV-2 B.1.351 variant after dose 2; further studies evaluating variant-specific modifications of mRNA-1283 as a booster dose are currently underway (NCT05137236) [28].

This study was strengthened by its randomized design, which allowed for descriptive comparisons to mRNA-1273, a widely administered SARS-CoV-2 vaccine with known efficacy in preventing COVID-19. A limitation was the small number of participants enrolled. This interim report presents findings through day 57 of an ongoing phase I trial that will continue to evaluate mRNA-1283 safety and immunogenicity, including long-term antibody responses to inform on immune response durability, which remains important amid reports of waning immunity within 6 months of mRNA-1273 primary vaccination [29, 30].

In conclusion, these preliminary findings support the continued evaluation of mRNA-1283, a next-generation SARS-CoV-2 vaccine. All dose levels of mRNA-1283 administered via a 2-dose regimen, 28 days apart, were generally safe in healthy adults aged 18-55 years, with the lowest dose level (10 μg) inducing comparable immunogenicity to the approved mRNA-1273 100 μg regimen. Clinical evaluation of mRNA-1283 is ongoing to assess the applicability of booster doses at lower dose levels and immune responses against other SARS-CoV-2 variants.

## Data Availability

Upon request, and subject to review, Moderna, Inc. will provide the data that support the findings of this study. Subject to certain criteria, conditions, and exceptions, Moderna, Inc. may also provide access to the related individual anonymized participant data.

## Acknowledgments

The authors would like to acknowledge Dr. Murray Kimmel (Optimal Research, Melbourne, Florida, USA), Dr. Daniel Brune (Optimal Research, Peoria, Illinois, USA), and Dr. Randall Severance (AES/Optimal Research, Chandler, Arizona, USA) for their contributions to the study. Medical writing and editorial assistance were provided by Emily Stackpole, PhD, of MEDiSTRAVA in accordance with Good Publication Practice (GPP3) guidelines, funded by Moderna, Inc., and under the direction of the authors. Potential conflictions of Interest: Y. D. P., L. S., A. A., U. S., and R. P. are employees of and shareholders in Moderna, Inc. P. Y. and M. H. have no conflicts to declare.

