## Supplementary Materials for "Interim Analysis of a Phase I Randomized Clinical Trial on the Safety and Immunogenicity of the mRNA-1283 SARS-CoV-2 Vaccine in Adults"

### Methods

#### *Inclusion and Exclusion Criteria*

Each participant was required to have met the following criteria for study enrollment:

- Male or female, 18 to 55 years of age at the time of consent
- Understood and agreed to comply with the study procedures and provided written informed consent
- In good general health and could comply with study procedures (based on investigator assessment)
- Body mass index of 18 kg/$m^2$ to 35 kg/$m^2$ (inclusive) at screening
- Had clinical screening laboratory evaluations (total white blood cell count, prothrombin time/partial thromboplastin time, hemoglobin, platelets, alanine aminotransferase, aspartate aminotransferase, creatinine, alkaline phosphatase, and total bilirubin) that were within normal reference ranges at the study-designated laboratory being used. Participants with grade 1 laboratory results at screening not considered clinically significant by the investigator could be re-screened once by assigning the participant a new identification number and repeating all screening procedures
- Female participants not of childbearing potential
- Female participants of childbearing potential could be enrolled in the study if fulfilling all of the following criteria:
    - Had a negative pregnancy test at screening and on the day of the first dose (day 1)
    - Had practiced adequate contraception or had abstained from all activities that could result in pregnancy for ≥28 days prior to the first dose (day 1)
    - Had agreed to continue adequate contraception through 3 months following the second dose (day 29)

  - Was not currently breastfeeding

Participants who met any of the following criteria were excluded from the study:

- Known history of severe acute respiratory syndrome coronavirus-2 (SARS-CoV-2) infection or known exposure to someone with SARS-CoV-2 infection or coronavirus disease 2019 (COVID-19) in the past 30 days
- Positive serology results for SARS-CoV-2 at screening
- Traveled outside of the United States in the 28 days prior to screening
- Was pregnant or breastfeeding at the time of screening or was planning to breastfeed at any time while enrolled in the study
- Was acutely ill or febrile 72 hours prior to or at screening or day 1. Fever was defined as a body temperature of ≥38.0°C/100.4°F. Participants meeting this criterion could be rescheduled within the 28-day screening window and would retain their initially assigned participant number. Afebrile participants with minor illnesses could be enrolled at the discretion of the investigator
- Prior administration of an investigational, authorized, or licensed CoV (eg, SARS-CoV-2, SARS-CoV, or Middle East respiratory syndrome coronavirus [MERS]-CoV) vaccine, based on medical history interview
- Current treatment with investigational agents for prophylaxis against COVID-19
- Recent (within the last 12 months) use of a dermal filler
- Was a healthcare worker or a member of an emergency response team
- Currently had symptomatic acute or chronic illness requiring medical or surgical care, to include changes in medication in the past 2 months indicating that chronic illness/disease was not stable (at the discretion of the investigator); this included any current workup of undiagnosed illness that could lead to a new condition

- Chronic disease to include: (a) uncontrolled hypertension based on age or systolic blood pressure of >150 mm Hg or diastolic blood pressure of >90 mm Hg at screening; (b) congestive heart failure; (c) unstable angina or exacerbation of coronary artery disease within 6 months before first study vaccination requiring cardiac intervention or new cardiac medications to control symptoms; (d) diabetes requiring use of medicine (insulin or oral) or not controlled with diet; or (e) chronic obstructive pulmonary disease, asthma requiring daily use of a bronchodilator or inhaled or systemic corticosteroids, or other chronic lung disease such as idiopathic pulmonary fibrosis
- Had a medical, psychiatric, or occupational condition that could pose additional risk as a result of participation, or that could interfere with safety assessments or interpretation of results according to the investigator's judgment
- Current or previous diagnosis of immunocompromising condition, immune-mediated disease, or other immunosuppressive condition
- Had received systemic immunosuppressants or immune-modifying drugs for >14 days in total within 6 months prior to screening (for corticosteroids ≥10 mg/day of prednisone equivalent) or was anticipating the need for immunosuppressive treatment at any time during participation in the study
- History of anaphylaxis, urticaria, or other significant adverse reaction requiring medical intervention after receipt of a vaccine
- Coagulopathy or bleeding disorder considered a contraindication to intramuscular injection or phlebotomy
- Had received or planned to receive any licensed vaccine ≤28 days prior to the first injection (day 1) or planned to receive a licensed vaccine within 28 days before or after any study injection, with the exception of licensed influenza vaccines, which may be received >14 days before the first study injection or >14 days after the second study injection

- Receipt of systemic immunoglobulins or blood products within 3 months prior to screening or plans for receipt during the study
- Current use of any inhaled substance (eg, tobacco or cannabis smoke, nicotine vapors)
- History of chronic smoking (≥1 cigarette a day) within 1 year of screening
- Resided in a nursing home
- Positive serology for hepatitis B virus surface antigen, hepatitis C virus antibody, or human immunodeficiency virus type 1 or 2 antibodies identified at screening
- Diagnosis of malignancy within previous 10 years (excluding non-melanoma skin cancer)
- Had donated ≥450 mL of blood products within 28 days prior to screening or had plans to donate blood products during the study
- Participated in an interventional clinical study within 28 days prior to screening based on the medical history interview or planned to do so while participating in this study
- Was an immediate family member or household member of study personnel, study site staff, or Sponsor personnel

***Randomization and Blinding***

This was an observer-blind study as to which vaccine or dose level was administered. All participants, study staff involved in participant assessment, and sponsor personnel were blinded to the individual dosing assignment until the end of the study. An unblinded staff member with no role in the observation or assessment of study participants prepared vaccines for administration. A limited number of sponsor and/or study personnel were unblinded to conduct safety data analyses for data and safety monitoring board safety data reviews and to perform the interim analysis.

#### *Clinical Laboratory Tests*

Blood samples for clinical laboratory tests were collected during screening and on days 8, 29, and 36. Clinical laboratory tests included hematology (hemoglobin, platelet count, and total white blood cell count), serum chemistry (alanine aminotransferase, aspartate aminotransferase, total bilirubin, alkaline phosphatase, and blood urea nitrogen), coagulation (prothrombin time and partial thromboplastin time), serology (hepatitis B surface antigen, hepatitis C virus antibody, and human immunodeficiency virus types 1 and 2 [screening only]), and other baseline screening analyses (urine pregnancy test [female participants of childbearing potential only] and urine follicle-stimulating hormone test [female participants to confirm postmenopausal status]).

#### *Recombinant Lentiviral-based Pseudovirus Neutralization Assay (Clinical Grade)*

As described previously [1], SARS-CoV-2 specific neutralizing antibodies were determined using lentivirus particles expressing the S protein of SARS-CoV-2 (Wuhan-Hu-1 with D614G mutation) or the B.1.351 variant (L18F-D80A-D215G-ΔL242-ΔA243-ΔL244-K417N-E484K-N501Y-D614G-A701V) on their surface as well as express the firefly luciferase reporter gene to quantitate infection by relative luminescence units (RLUs). Pseudoviruses were applied to 293T cells stably transduced to express high levels of angiotensin-converting enzyme 2 (ACE2) with or without antibody pre-incubation (control antibodies or serum samples). Dose-response curves were developed using serial dilution of control antibodies or serum samples; neutralization was measured as the serum dilution whereby RLUs were reduced by 50% (50% inhibitory dilution; $ID_{50}$) relative to the mean RLUs for the virus control (cells and virus without control antibody or serum samples) after subtraction of the mean RLUs in the cell control (cells only).

***SARS-CoV-2 Microneutralization Assay***

This assay followed a previously published protocol [2]. Serum neutralizing antibodies against SARS-CoV-2 were quantified via an *in situ* enzyme linked immunosorbent assay (ELISA). This SARS-CoV-2 microneutralization assay is a validated assay for quantitation of neutralizing antibodies to SARS-CoV-2 in human serum.

Vero E6 cells were seeded in 96-well tissue culture plates and maintained at 37 ± 2 °C and 5.0 ± 2% $CO_2$ until 90-100% monolayer confluency. The sample dilution series of heat-inactivated serum samples (30 ± 5 minutes at 56.0 °C) was performed in titer tubes with the initial dilution, followed by 7 serial 2-fold dilutions. SARS-CoV-2 Wuhan-Hu-1 (without D614G mutation) viral stock was thawed at 37.0 °C and kept on ice prior to diluting in sample diluent (Eagle's minimal essential media with 1 mM sodium pyruvate, 2 mM L-glutamine, 1% volume/volume [v/v] penicillin-streptomycin, and 5% v/v heat-inactivated fetal bovine serum. The virus stock was prepared at a target concentration of 200 median tissue culture infectious doses ($TCID_{50}$) per well (2000 $TCID_{50}$/mL) and added to each sample dilution in equal volumes. Virus-serum mixtures were incubated for 60 ± 5 minutes at 37.0 ± 2 °C and 5.0 ± 2% $CO_2$ and then added to cell monolayers followed by incubation for 40-46 hours at 37.0 ± 2 °C and 5.0 ± 2% $CO_2$. After the incubation, the inoculum was removed and cells were washed with Hank's Balanced Salt Solution then fixed with 80% cold acetone for at least 30 minutes. Fixed cells were incubated with anti-nucleocapsid protein primary antibody cocktail (clones HM1056 and HM1057; EastCoast Bio, North Berwick, ME) for 60 ± 5 minutes at 37.0 ± 2 °C, washed, then incubated with goat anti-mouse IgG Horse Radish Peroxidase (HRP) conjugate (Fitzgerald, North Acton, MA) for 60 ± 5 minutes at 37.0 ± 2 °C. Plates were washed and ABTS Microwell Peroxidase Substrate System (Kirkegaard and Perry [KPL]; KPL #50-62-00) added to each well. After a short incubation, ABTS Peroxidase Stop Solution (KPL #50-85-01) was added to stop the reaction and the optical density was measured at 405 nm using a 490 nm reference wavelength. Each well was scored as positive or negative for viral infection based on the plate cutoff value. Neutralization was defined as a ≥50% reduction of the viral challenge signal via the following equation: ([average of the virus control wells – average of the cell control wells]/2) + average of the cell control wells.

#### *SARS-CoV-2 Meso Scale Discovery 4-PLEX Immunoassay*

This quantitative electrochemiluminescence (ECL) method is an indirect binding ECL method used to detect SARS-CoV-2 antibodies in human serum. Based on Meso Scale Discovery (MSD) technology, the assay employs capture molecule MultiSPOT microtiter plates fitted with a series of electrodes. Specifically, a MSD MESO SECTOR S 600 detection system was used to apply an electrical current to custom microtiter plates that led to light emission via SULFO-TAG™ through a series of oxidation-reduction reactions with ruthenium and tripropylamine (TPA), which was quantitated using a plate reader measuring the intensity of emitted light. The bio-assay utilized a ten-spot custom SARS-CoV-2 4-PLEX (modified from a previously described 3-PLEX assay [3]) plate coated with SARS-CoV-2 antigens specific to the full length S protein (Wuhan-Hu-1 isolate with D614G mutation), nucleocapsid, and N-terminal domain (NTD) and receptor-binding domain (RBD). Anti-SARS-CoV-2 antibodies present in samples that bound to the antigen-coated plates and formed antibody-antigen complexes were detected with the addition of SULFO-TAG™-labeled antibodies. TPA buffer solution was added to result in ECL that was measured in RLU with the MSD SECTOR S 600 plate reader. Antibody concentrations were quantitated by interpolating ECL response based on the standard curve generated from a serially diluted reference standard.

#### *Analysis Populations and Sample Size*

The safety population constituted all randomly assigned participants who received ≥1 dose of the study vaccination while the solicited safety population consisted of all randomly assigned participants receiving ≥1 dose and contributing any solicited AR data. The per-protocol population included all randomly assigned participants who received ≥1 dose of the study vaccination and had no major protocol deviations that affected immunogenicity data, did not have SARS-CoV-2 infection, complied with the vaccination schedule, and complied with the timing for blood sampling to have post-vaccination immunogenicity results for ≥1 assay component of the immunogenicity objective

The planned sample size provided a >90% probability to observe ≥1 participant with an AE if the true rate of AEs was 3%. The sample size for the trial was not driven by statistical assumptions for formal hypothesis testing. No formal statistical hypotheses were tested in this study.

**Results**

***Clinical Laboratory Tests***

The most common laboratory abnormality was decreased hemoglobin level from baseline, which was primarily grade 1 or grade 2 and reported by 70.0-76.2% of participants across mRNA-1283 groups and 68.2% of mRNA-1273 recipients after dose 1 (day 8). After dose 2 (day 36), decreased hemoglobulin level was reported by 47.6-80.0% of participants across mRNA-1283 groups and by 60.0% of recipients of mRNA-1273. Increased alanine aminotransferase level (grade 1 or 2) was reported for 6 participants after dose 1 (day 8; mRNA-1283 10 µg, n=2 [10.0%]; mRNA-1283 30 µg, n=1 [4.8%]; mRNA-1283 100 µg, n=1 [4.8%]; mRNA-1273, n=2 [9.1%]) and 4 participants after dose 2 (day 36; mRNA-1283 10 µg, n=2 [10.5%]; mRNA-1283 100 µg, n=2 [9.5%]). Increased aspartate aminotransferase level (primarily grade 1) after dose 1 was reported for 0-5.0% of participants across mRNA-1283 groups and 4.5% of recipients of mRNA-1273; increased level occurred in 1 participant in the mRNA-1283 10-µg group after dose 2. Four participants reported abnormal laboratory values as treatment-emergent adverse events: decreased hemoglobin level (mRNA-1283 100 µg, n=2; mRNA-1273 100 µg, n=1) and increased transaminase levels (mRNA-1273 100 µg, n=1); none were considered serious and 2 were considered treatment-related by the investigator (mRNA-1283 100 µg, n=1 [decreased hemoglobin level]; mRNA-1273 100 µg, n=1 [increased transaminase levels]).

**Supplementary Table 1. Summary of neutralizing antibody responses against SARS-CoV-2 at day 57 (per-protocol immunogenicity population)**

| | mRNA-1283 | | | | mRNA-1273 |
|---|---|---|---|---|---|
| **Time point**<br>**Analysis** | **10 µg (n = 16)** | **30 µg (n = 18)** | **100 µg (n = 20)** | **Placebo + 100 µg (n = 14)** | **100 µg (n = 18)** |
| Pseudovirus neutralizing antibody $ID_{50}$ titers against D614G SARS-CoV-2 | | | | | |
| **Baseline (day 1)** | | | | | |
| n | 16 | 18 | 20 | 14 | 18 |
| GMT (95% CI) | 9.3 (NE) | 12.1 (8.0-18.1) | 9.3 (NE) | 14.6 (5.5-39.0) | 9.3 (NE) |
| Number of participants ≥ LLOQ, n | 0 | 2 | 0 | 1 | 0 |
| **Month 2 (day 57)** | | | | | |
| n[a] | 16 | 18 | 20 | 14 | 18 |
| GMT (95% CI) | 1370 (888-2112) | 1091 (645-1846) | 1679 (1133-2487) | 58.6 (16.6-206.0) | 1190 (826-1713) |
| GMR[b] (95% CI) | 1.15 (0.67-1.98) | 0.92 (0.50-1.70) | 1.41 (0.84-2.38) | 0.05 (0.01-0.18) | NA |
| GMFR[c] (95% CI) | 148.1 (96.0-228.3) | 90.4 (48.0-170.0) | 181.5 (122.5-268.9) | 4.02 (1.52-10.6) | 128.6 (89.3-185.2) |
| Seroresponse,[d] n (%) [95% CI] | 16 (100.0) [79.4-100.0] | 18 (100.0) [81.5-100.0] | 20 (100.0) [83.2-100.0] | 4 (28.6) [8.4-58.1] | 18 (100.0) [81.5-100.0] |
| Live virus neutralizing antibody 50% microneutralization against Wuhan-Hu-1 (without D614G mutation) | | | | | |
| **Baseline (day 1)** | | | | | |
| n | 16 | 18 | 20 | 14 | 18 |
| GMT (95% CI) | 79.9 (NE) | 79.9 (NE) | 79.9 (NE) | 115.8 (51.9-258.4) | 79.9 (NE) |
| Number of participants ≥ LLOQ, n | 0 | 0 | 0 | 1 | 0 |

| | | | | | |
|---|---|---|---|---|---|
| **Month 2 (day 57)** | | | | | |
| n[a] | 16 | 18 | 20 | 14 | 18 |
| GMT (95% CI) | 2803 (1801-4364) | 2253 (1448-3505) | 4371 (3161-6043) | 204 (73-571) | 2950 (2335-3726) |
| GMR[b] (95% CI) | 0.95 (0.58-1.55) | 0.76 (0.47-1.24) | 1.48 (1.00-2.20) | 0.07 (0.02-0.20) | NA |
| GMFR[c] (95% CI) | 35.1 (22.5-54.6) | 28.2 (18.1-43.9) | 54.7 (39.6-75.6) | 1.8 (0.9-3.4) | 36.9 (29.2-46.6) |
| Seroresponse,[d] n (%) [95% CI] | 15 (93.8) [69.8-99.8] | 16 (88.9) [65.3-98.6] | 20 (100.0) [83.2-100.0] | 1 (7.1) [0.2-33.9] | 18 (100.0) [81.5-100.0] |
| Pseudovirus neutralizing antibody $ID_{50}$ titers against B.1.351 | | | | | |
| **Baseline (day 1)** | | | | | |
| n | 16 | 18 | 20 | 14 | 18 |
| GMT (95% CI) | 9.8 (NE) | 13.2 (7.8, 22.1) | 9.8 (NE) | 14.0 (6.4, 30.6) | 9.8 (NE) |
| Number of participants ≥ LLOQ, n | 0 | 2 | 0 | 1 | 0 |
| **Month 2 (day 57)** | | | | | |
| n[a] | 16 | 18 | 20 | 14 | 18 |
| GMT (95% CI) | 138.0 (83.6-227.9) | 113.0 (61.4-208.1) | 144.6 (89.6-233.5) | 25.2 (8.3-75.9) | 107.7 (78.4-148.0) |
| GMR[b] (95% CI) | 1.28 (0.74-2.24) | 1.05 (0.54-2.05) | 1.34 (0.76-2.37) | 0.23 (0.08-0.73) | NA |
| GMFR[c] [95% CI] | 14.2 (8.6-23.4) | 8.59 (4.2-17.4) | 14.8 (9.2-23.9) | 1.80 (0.7-4.8) | 11.0 (8.0-15.2) |
| Seroresponse,[d] n (%) [95% CI] | 12 (75.0) [47.6-92.7] | 11 (61.1) [35.7-82.7] | 17 (85.0) [62.1-96.8] | 2 (14.3) [1.8-42.8] | 12 (66.7) [41.0-86.7] |

The LLOQ was 18.5 for the pseudovirus neutralizing antibody $ID_{50}$ titers and 19.5 for the pseudovirus neutralizing antibody $ID_{50}$ titers against B.1.351. Numbers below the LLOQ were replaced by 0.5 x LLOQ. One participant in placebo + mRNA-1283 100 µg and 2 participants in the mRNA-1283 30 µg group had pseudovius neutralizing antibodies to Wuhan-Hu-1 (with D614G mutation) SARS-CoV-2 and B.1.351 by that were above the LLOQ at baseline, suggestive of previous infection despite negative SARS-CoV-2 serology at baseline.

Abbreviations: CI, confidence interval; GMFR, geometric mean fold rise; GMR, geometric mean ratio; GMT, geometric mean titer; LLOQ, lower limit of quantification; mRNA, messenger RNA; NA, not applicable; NE, not estimable; SARS-CoV-2, severe acute respiratory syndrome coronavirus-2.

[a]Number of participants in the per-protocol immunogenicity population with non-missing data at that time point.

[b]GMR of mRNA-1283 versus mRNA-1273.

[c]GMFR of titers from baseline to postbaseline time point.

[d]Seroresponse was defined as an increase from below the LLOQ to ≥4 x LLOQ if baseline antibody level was <LLOQ, or an ≥4-fold increase if baseline antibody level was ≥LLOQ.

**Supplementary Table 2. Summary of binding antibody responses against SARS-CoV-2 at day 57 (per-protocol immunogenicity population)**

| | mRNA-1283 | | | | mRNA-1273 |
|---|---|---|---|---|---|
| **Timepoint**<br>**Analysis** | **10 µg**<br>**(n = 16)** | **30 µg**<br>**(n = 18)** | **100 µg**<br>**(n = 20)** | **Placebo +**<br>**100 µg (n = 14)** | **100 µg**<br>**(n = 18)** |
| SARS-CoV-2 S protein IgG antibody (AU/mL) by MSD 4-PLEX assay | | | | | |
| **Baseline (day 1)** | | | | | |
| n | 16 | 18 | 20 | 14 | 18 |
| GM level (95% CI) | 46.8<br>(18.8-116.5) | 52.0<br>(20.9-129.5) | 42.9<br>(21.3-86.7) | 79.4<br>(14.9-423.0) | 42.4<br>(26.4-68.0) |
| Number of participants ≥ LLOQ, n | 10 | 13 | 13 | 10 | 15 |
| **Month 2 (day 57)** | | | | | |
| n[a] | 16 | 18 | 20 | 14 | 18 |
| GM level (95% CI) | 322,671<br>(232,477-<br>447,858) | 280,454<br>(196,077-<br>401,139) | 416,620<br>(312,082-<br>556,175) | 15,396<br>(4855-48,820) | 303,004<br>(242,750-<br>378,212) |
| GMR[b] (95% CI) | 1.06<br>(0.73-1.55) | 0.93<br>(0.62-1.39) | 1.37<br>(0.96-1.97) | 0.05<br>(0.02-0.16) | NA |
| GMFR[c] [95% CI] | 6894<br>(2646-17,964) | 5394<br>(2012-14,457) | 9702<br>(4746-19,833) | 193.9<br>(56-674) | 7146<br>(3891-13,126) |
| Seroresponse,[d] n (%) [95% CI] | 16 (100.0)<br>[79.4-100.0] | 18 (100.0)<br>[81.5-100.0] | 20 (100.0)<br>[83.2-100.0] | 13 (92.9)<br>[66.1-99.8] | 18 (100.0)<br>[81.5-100.0] |
| SARS-CoV-2 RBD IgG antibody (AU/mL) by MSD 4-PLEX assay | | | | | |
| **Baseline (day 1)** | | | | | |
| n | 16 | 18 | 20 | 14 | 18 |
| GM level (95% CI) | 34.1<br>(15.1-77.3) | 25.3<br>(14.6-43.8) | 17.2<br>(12.0-24.6) | 39.9<br>(7.5-213.3) | 21.2<br>(13.0-34.5) |
| Number of participants ≥ LLOQ, n | 9 | 11 | 9 | 5 | 9 |

| | | | | | |
|---|---|---|---|---|---|
| **Month 2 (day 57)** | | | | | |
| n[a] | 16 | 18 | 20 | 14 | 18 |
| GM level (95% CI) | 231,321 (162,244-329,808) | 204,219 (144,945-287,734) | 297,202 (216,874-407,285) | 9700 (3110-30,252) | 154,862 (122,853-195,211) |
| GMR[b] (95% CI) | 1.49 (1.00-2.22) | 1.32 (0.89-1.96) | 1.92 (1.31-2.82) | 0.06 (0.02, 0.20) | NA |
| GMFR[c] (95% CI) | 6779 (2686-17,110) | 8086 (3917-16,692) | 17,333 (10,881-27,609) | 243.1 (81-726) | 7321 (4042-13,261) |
| Seroresponse,[d] n (%) [95% CI] | 16 (100.0) [79.4-100.0] | 18 (100.0) [81.5-100.0] | 20 (100.0) [83.2-100.0] | 13 (92.9) [66.1-99.8] | 18 (100.0) [81.5-100.0] |
| SARS-CoV-2 NTD IgG antibody (AU/mL) by MSD 4-PLEX assay | | | | | |
| **Baseline (day 1)** | | | | | |
| n | 16 | 18 | 20 | 14 | 18 |
| GM level (95% CI) | 2.6 (1.6-4.2) | 2.3 (1.5-3.6) | 1.8 (1.4-2.3) | 4.5 (1.0-20.4) | 2.0 (1.5-2.6) |
| Number of participants ≥ LLOQ, n | 5 | 5 | 3 | 4 | 4 |
| **Month 2 (day 57)** | | | | | |
| n[a] | 16 | 18 | 20 | 14 | 18 |
| GM level (95% CI) | 5957 (4230-8390) | 5401 (3467-8415) | 8775 (6375-12,078) | 296 (95, 919) | 5584 (4501, 6928) |
| GMR[b] (95% CI) | 1.07 (0.73-1.56) | 0.97 (0.60-1.57) | 1.57 (1.07-2.30) | 0.05 (0.02-0.17) | NA |
| GMFR[c] (95% CI) | 2326 (1221-4432) | 2341 (1109-4942) | 4849 (3230-7280) | 65 (26-164) | 2855 (1895-4303) |
| Seroresponse,[d] n (%) [95% CI] | 16 (100.0) [79.4-100.0] | 18 (100.0) [81.5-100.0] | 20 (100.0) [83.2-100.0] | 13 (92.9) [66.1-99.8] | 18 (100) [81.5-100.0] |

The LLOQ was 23 for the SARS-CoV-2 S protein IgG antibody, 19 for the SARS-CoV-2 RBD IgG antibody, and 3 for the SARS-CoV-2 NTD IgG antibody. Numbers below the LLOQ were replaced by 0.5 x LLOQ.

Abbreviations: CI, confidence interval; GM, geometric mean; GMFR, geometric mean fold rise; GMR, geometric mean ratio; IgG, immunoglobulin G; LLOQ, lower limit of quantification; mRNA, messenger RNA; MSD, Meso Scale Discovery; NA, not applicable; NTD, N-terminal domain; RBD, receptor binding domain; S, Spike; SARS-CoV-2, severe acute respiratory syndrome coronavirus-2.

[a]Number of participants in the per-protocol immunogenicity population with non-missing data at that time point..

[b]GMR of mRNA-1283 versus mRNA-1273.

[c]GMFR of levels from baseline to postbaseline time point.

[d]Seroresponse was defined as an increase from below the LLOQ to ≥4 x LLOQ if baseline antibody level was <LLOQ, or an ≥4-fold increase if baseline antibody level was ≥LLOQ.

**Supplementary Figure 1. Study design.**

A total of 104 adult participants were randomly assigned (1:1:1:1:1) in parallel to each group.

Participants were randomly assigned to receive 2 doses of mRNA-1283 (10 µg, 30 µg, or 100 µg) or mRNA-1273 (100 µg) administered 28 days apart (day 1 and day 29) or to receive 1 dose of mRNA-1283 (100 µg) on day 29 with placebo administered on day 1. This report summarizes interim safety and immunogenicity findings through day 57.

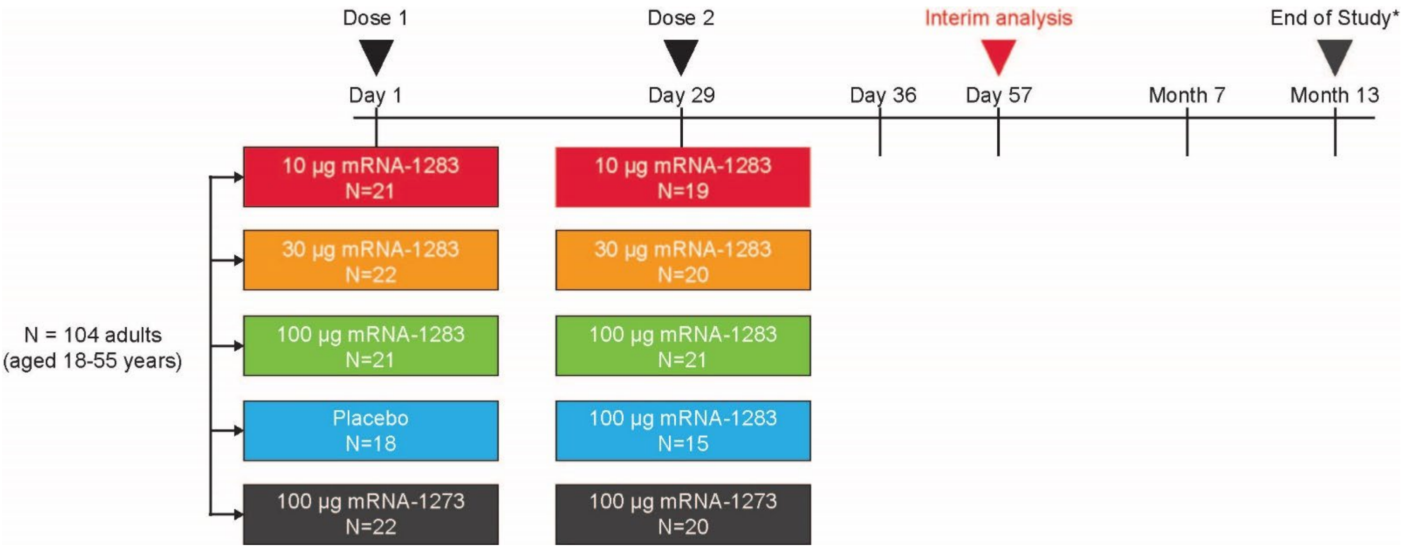


*Participants will have the option to continue follow-up visits at month 19 and month 25.

Abbreviation: mRNA, messenger RNA.

**Supplementary Figure 2. Wuhan-Hu-1 (without D614G mutation) live virus neutralizing antibody 50% microneutralization titers.** Neutralizing antibody titers on the microneutralization assay at 50% are shown for those participants in the per-protocol immunogenicity population. Boxplots are based on log-transformed values; boxes and horizontal bars denote IQR range and $ID_{50}$, respectively, with whisker end points equal to the values below or above the median at 1.5 times the IQR. Dots represent individual results and the dotted lines represent either the LLOQ or ULOQ. Antibody values reported as below the LLOQ were replaced by 0.5 x LLOQ, whereas values greater than the ULOQ were converted to the ULOQ if actual values were not available. The LLOQ was 159.79 ($log_{10}$[LLOQ] = 2.204) and the ULOQ was 11173.11 ($log_{10}$[ULOQ] = 4.048).

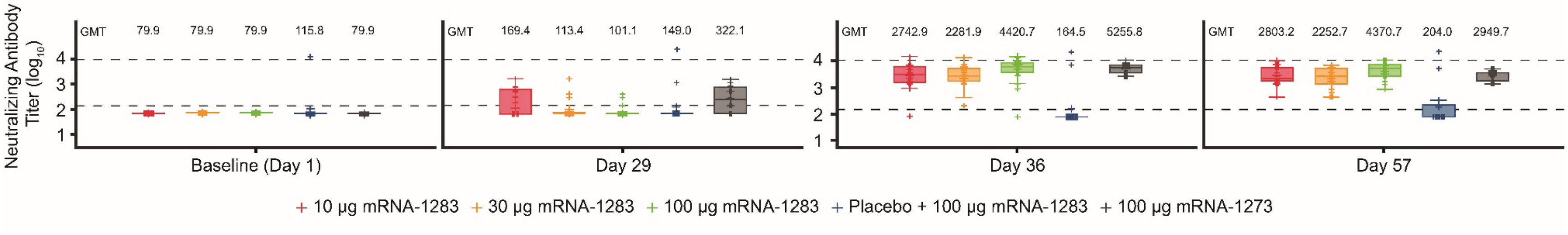


Abbreviations: $ID_{50}$, 50% inhibitory dose; IQR, interquartile range; LLOQ, lower limit of quantification; mRNA, messenger RNA; ULOQ, upper limit of quantification.
